# The Social Determinants of Depression among Adolescents in Low and Middle Income Countries: A Scoping Review Protocol

**DOI:** 10.1101/2024.10.04.24314891

**Authors:** Isayas Wubshet, Kibur Engedawor, Semere Gebremariam, Clementine Kanazayire, Pamela Abbott

## Abstract

**Background:** Adolescence is a critical and stressful period characterized by rapid biological, psychological, and social changes. Early signs of depression often emerge during early adolescence. Apart from biological and psychological factors, the interplay among different social factors in the family, school, and neighborhood plays a significant role in determining depression among adolescents. Although depression is highly prevalent in low and middle-income countries, there is a shortage of comprehensive scoping reviews that examine the social factors and their relationships.

**Objective:** the purpose of this review is to gain an understanding of the social determinants of adolescent depression in family, school, and neighborhood settings.

**Method and analysis:** This protocol will adhere to Arksey and O’Malley’s framework for conducting scoping reviews, meticulously detailing each stage of the process. Firstly, we will use predetermined keywords to systematically search across eight electronic resources. Two reviewers will then carefully assess the titles and abstracts of retrieved studies. Only those deemed pertinent will proceed to full-text screening. Studies will be selected according to established inclusion and exclusion criteria. The review team will conduct the data extraction procedure on its own. Finally, the included studies will undergo thorough qualitative and quantitative analyses. The results will be presented at conferences and published in peer-reviewed journals.

## 1. Background

Good mental wellbeing relies on healthy psychological, biological, and social environments. Distortions in these environments can detrimentally affect mental health, contributing to various mental illnesses. Globally, approximately 970 million individuals suffer from mental health disorders, with depression ranking the third leading burden of disease worldwide (WHO, 2018b). Between 2005 and 2015, there was an 18.4% increase in the number of persons living with depression (Vos et al., 2016), reaching 300 million in 2018 (WHO, 2018b). By 2030, depression is expected to rank second in terms of causes of disability, excluding the 25% rise linked to COVID-19 (Rehm & Shield, 2019; WHO, 2022).

Depression affects the healthy functioning of a person (Petersen et al., 1993). It has physiological and socio-emotional manifestations such as loss of appetite, tiredness, low mood, sadness, a feeling of guilt (Wang et al., 2016), feelings of inferiority (Weersing et al., 2016; Zhu & Wong, 2022), and personal and social life displeasure, which affect anyone regardless of gender and age (Schaan, 2013). When these symptoms persist, a person loses interest in activities they usually like and becomes unable to perform everyday tasks (WHO, 2017).

Early depressive signs begin to manifest at a younger age (Abela & Hankin, 2008; Hankin, 2006; Jenness et al., 2019; Upashree D. & Maraichelvi, 2020). WHO defines adolescence as the life span from 10 to 19 years (WHO, 2021), which is a transitional stage from childhood to adulthood that results in physical, psycho-emotional, and social changes (Alshammari et al., 2021; Ibrahim et al., 2017; Jayanthi et al., 2015; Sreekanth et al., 2023). This age group is considered “critical” (Mukangabire et al., 2021), and this is primarily because the rapid development observed during adolescence exposes them to many risky behaviors that affect their current and future lives (Hunduma et al., 2022; Mekdes et al., 2018). Adolescence is a period of uncertainty (Ibrahim et al., 2017), vulnerability, risk (De-la-Iglesia & Olivar, 2015), taking responsibility (Tirfeneh & Srahbzu, 2020), and problems with relationships (Frojd et al., 2008).

A WHO report (2019) disclosed that depression is a significant mental disorder among adolescents, accounting for 16% of the mental disorder burden and leading to 40-60% premature mortality (Minichil et al., 2019). Shefaly et al.’s (2022) research reveals that 34% of adolescents are vulnerable to depression, with the rate surpassing 18-25 years, leading to one million avoidable adolescent deaths.

Eighty percent of adolescents who have experienced a mental condition in their lifetime report having depression. These adolescents reside in low- and middle-income countries (LMICs), home to around ninety percent of the 1.2 billion adolescents worldwide (UN, as referenced in Jason et al., 2018). This pattern is remarkably noticeable in African and Asian nations, where communicable diseases are more prevalent. According to Presentati (2021), adolescents in sub-Saharan Africa have a 26.9% prevalence of depression.

Adolescent depression is no longer an individual issue; rather, it is now a social pathology that causes both immediate and long-term societal difficulties and crises. Adolescents participate in a range of social interactions and engage in various social networks, which can significantly impact their mental well-being, either positively or negatively. Therefore, attempting to view adolescents independently of these social contexts is akin to separating the soul from the body, rendering both devoid of life. Thus, it is crucial to examine the significant external social factors that extend beyond the examination of an individual’s internal bio-psychological factors (Muntaner et al., 2013; Turner et al., 2014).

While etiologically based scoping reviews on adolescent depression, particularly focusing on social factors, remain limited, the research team is deeply concerned by the absence of previous studies addressing low- and middle-income countries. Conversely, emerging research highlights a troubling surge in adolescent depression within these countries, imposing a mounting public health burden. Therefore, understanding the complexities of external socio-structural factors within family, school, and community settings, which wield significant influence over the outcomes of adolescent depressive symptoms, becomes imperative for a comprehensive understanding of the issue and for effective service delivery. Thus, the purpose of this scoping review protocol is to provide a guide on how to undertake a scoping review of the social contexts that lead to and prevent adolescent depression in low and middle-income countries.

## 2. Method

The primary goal of this protocol is to detail the approach for carrying out a scoping review on the social factors that affect depression in adolescents living in low- and middle-income countries. It will follow the structure of a scoping review introduced by Arksey and O’Malley (2005), which includes the following steps: identifying the research question, searching for relevant studies, selecting studies, mapping out the data, summarizing and reporting the results, and including expert consultation. Any modifications or improvements made throughout the review will be integrated into the final scoping review paper.

### 2.1. Identifying the research question

Drawing from a preliminary exploration of the literature, the research questions have been formulated. The primary research question of this review is to identify the major social risk and protective factors contributing to the development of depression among adolescents (aged 10-18 years) in low- and middle-income countries. Following the primary research question, the specific research objectives are outlined as follows:

- What are the differences between boys and girls, as well as different socio-economic groups, in terms of depressive symptoms?
- In what ways do family contexts contribute to or prevent depressive symptoms among adolescents in LMICs?
- How do school environments influence the occurrence or prevention of depressive symptoms among adolescents in LMICs?
- What role do neighborhood contexts play in either fostering or mitigating depressive symptoms among adolescents in LMICs?
- How do social factors within family, school, and community settings interact to impact adolescent depressive symptoms in LMICs?

### 2.2. Identifying Relevant Studies

This review will encompass both peer-reviewed studies and gray literature. The electronic databases to be utilized include Google Scholar, PsychInfo (via Ovid), Web of Science, PubMed, Sociological Abstracts (via Proquest), and Embase (via Ovid). Moreover, the reference lists of the included studies will be carefully examined to identify any relevant literature.

Search terms will be employed to query all databases for literature published from January 2010 to December 2023. Literature written in languages other than English will not be included. The following table outlines the search strategy.

| S/N | Database | Search terms |  |  | Language | Date |
| --- | --- | --- | --- | --- | --- | --- |
|  |  | Age | Social determinants | Countries |  |  |
| 1 | Web of science | “adolescent”<br>OR “child”<br>OR “teenager”<br>OR “youth”<br>OR “young”<br>OR “juvenile”<br>OR “pediatric” | “academic performance” OR<br>“academic stress” OR “bullying”<br>OR “discrimination” OR “economic<br>inequality” OR “economic status”<br>OR “educational opportunities” OR<br>“family dynamics” OR “family<br>factors” OR “family process” OR<br>“family types” OR “financial stress”<br>OR “interpersonal relationships”<br>OR “neighborhood conditions” OR<br>“parental conflict” OR “peer<br>relationships” OR “poverty” OR<br>“racism” OR “school factors” OR<br>“social class” OR “social<br>consequences” OR “social<br>determinants” OR “social isolation”<br>OR “social network” OR “Social<br>relation” OR “socio-demographic”<br>OR “social status” OR “social<br>support” OR “socio-cultural<br>meaning” OR “socioeconomic<br>status” OR “traumatic life event”<br>OR “unemployment” OR “parental<br>mental disorder” OR “parenting”<br>OR “victim” OR “abuse” OR<br>“violence” OR “domestic violence”<br>OR “adverse childhood experience”<br>OR “social capital” OR<br>“maltreatment” OR “mental<br>treatment” OR “stressful event” OR<br>“social mediator” OR “social<br>intervention” OR “parental support”<br>OR “peer” OR “mental policy” OR<br>“mental treatment” OR “cultural<br>meaning” OR “Socio-cultural<br>factors” OR “sociocultural” | “low income” OR “middle income” OR “low and middle income”<br>OR “Afghanistan” OR “Burkina Faso” OR “Burundi” OR “Central<br>African Republic” OR “Chad” OR “Guinea-Bissau” OR “Korea”<br>OR “Liberia” OR “Madagascar” OR “Malawi” OR “Somalia” OR<br>“South Sudan” OR “Sudan” OR “Syria” OR “Togo” OR “Congo”<br>OR “Eritrea” OR “Ethiopia” OR “Gambia” OR “Guinea” OR<br>“Mali” OR “Mozambique” OR “Niger” OR “Rwanda” OR “Sierra<br>Leone” OR “Uganda” OR “Yemen” OR “Zambia” OR “Angola”<br>OR “Algeria” OR “Bangladesh” OR “Benin” OR “Bhutan” OR<br>“Bolivia” OR “Cape Verde” OR “Cambodia” OR “Cameroon” OR<br>“India” OR “Indonesia” OR “Iran” OR “Kenya” OR “Kiribati” OR<br>“Kyrgyzstan” OR “Lao” OR “Laos” OR “Lebanon” OR “Lesotho”<br>OR “Philippines” OR “Samoa” OR “Sao Tome and Principe” OR<br>“Senegal” OR “Solomon Islands” OR “Sri Lanka” OR “Tanzania”<br>OR “Tajikistan” OR “Timor-Leste” OR “Comoros” OR “Republic<br>of the Congo” OR “Cote d’Ivoire” OR “Djibouti” OR “Mauritania”<br>OR “Micronesia” OR “Mongolia” OR “Morocco” OR “Tunisia”<br>OR “Ukraine” OR “Uzbekistan” OR “Vanuatu” OR “Egypt” OR<br>“El Salvador” OR “Eswatini” OR “Ghana” OR “Haiti” OR<br>“Honduras” OR “Myanmar” OR “Nepal” OR “Nicaragua” OR<br>“Nigeria” OR “Pakistan” OR “Papua New Guinea” OR “Vietnam”<br>OR “West Bank and Gaza” OR “Zimbabwe” OR “Albania” OR<br>“American Samoa” OR “Argentina” OR “Armenia” OR<br>“Azerbaijan” OR “Belarus” OR “Belize” OR “Bosnia and<br>Herzegovina” OR “Fiji” OR “Gabon” OR “Georgia” OR<br>“Grenada” OR “Guatemala” OR “Guyana” OR “Iraq” OR<br>“Jamaica” OR “Namibia” OR “North Macedonia” OR “Palau” OR<br>“Paraguay” OR “Peru” OR “Russian Federation” OR “Russia” OR<br>“Serbia” OR “South Africa” OR “Botswana” OR “Jordan” OR “St<br>Lucia” OR “Brazil” OR “Bulgaria” OR “China” OR “Colombia”<br>OR “Costa Rica” OR “Cuba” OR “Dominica” OR “Dominican<br>Republic” OR “Equatorial Guinea” OR “Kazakhstan” OR<br>“Kosovo” OR “Libya” OR “Malaysia” OR “Maldives” OR<br>“Marshall Islands” OR “Mauritius” OR “Mexico” OR “Moldova”<br>OR “Vincent and the Grenadines” OR “Suriname” OR “Thailand”<br>OR “Tonga” OR “Turkiye” OR “Turkey” OR “Turkmenistan” OR<br>“Tuvalu” OR “Ecuador” OR “Montenegro” | English | January,<br>2010-December,<br>2023 |
| 2 | PubMed |  |  |  |  |  |
| 3 | Embase |  |  |  |  |  |
| 4 | Sociological<br>abstracts |  |  |  |  |  |
| 5 | Psychinfo |  |  |  |  |  |
| 6 | Google scholar |  |  |  |  |  |

### 2.3. Study Selection

The online scoping review tool Covidence will be used to help with the study selection process. Literature obtained from search engines for both peer-reviewed and gray literature will be exported to Endnote and subsequently imported to Covidence. Upon the removal of duplicate citations, the initial stage of screening will commence, where two reviewers will conduct the title and abstract screening for all studies retrieved by the search strategy. Any conflicts will be resolved by a third screener, who will engage in discussions with the other two screeners to reach an agreement.

After removing papers that do not meet the inclusion criteria based on their titles and abstracts, the next step involves carefully assessing the remaining records to establish their eligibility based on the established criteria. In addition to publications identified through refined database searches, those found manually will also be included.

A first independent screening of a sample of retrieved records will be carried out by two team members to guarantee consistent application of the eligibility criteria. Records identified as relevant by either reviewer will advance to the full-text review stage. Inter-rater reliability will be assessed using the percentage agreement, targeting a minimum threshold of 80% (McHugh, 2012). The reviewers will examine any differences in eligibility determinations and, if necessary, involve a third-party adjudicator to settle the issues.

In cases where a correction to an article has been made after publication, the corrected version will be used. The authors will be notified by email if any information is missing or if further explanation of the data is needed. Records not available electronically will be procured via interlibrary loan through the University of Aberdeen, with a 30-day allowance for receipt.

| Inclusion criteria | Exclusion criteria |
| --- | --- |
| Studies including adolescents aged 10-18 years. | Studies dealing with people below 10 years old and adult people (above the age of 18) |
| Studies on adolescents living in the community (such as with their families, relatives, so on) | Studies that only include adolescents in residential care, hospital foster care or in prison |
| Studies exhaustively or mainly on depression | Studies dealing with other mental and psycho-emotional health issues |
| Focusing on one or more social factors for adolescent depression | Studies focusing only on measuring prevalence and associated factors (the factors that are not exhaustively examined rather than simply showing the statistical significance) |
| Being a universal sample | Studies focused on specific medical cohorts<br>Natural disaster-related depression (e.g. due to earthquake, flood, etc) |
| Empirical studies | Opinion papers, letters to the editor, editorials, or comments |
|  | • Systematic and scoping review<br>• Studies for testing model, theory, approach, platform, intervention, tool |
| • Urban setting | • Non-urban and mixed (both urban and rural) setting |
| • Studies written in English language | • Studies written in other language than English |

**Fig 1.**
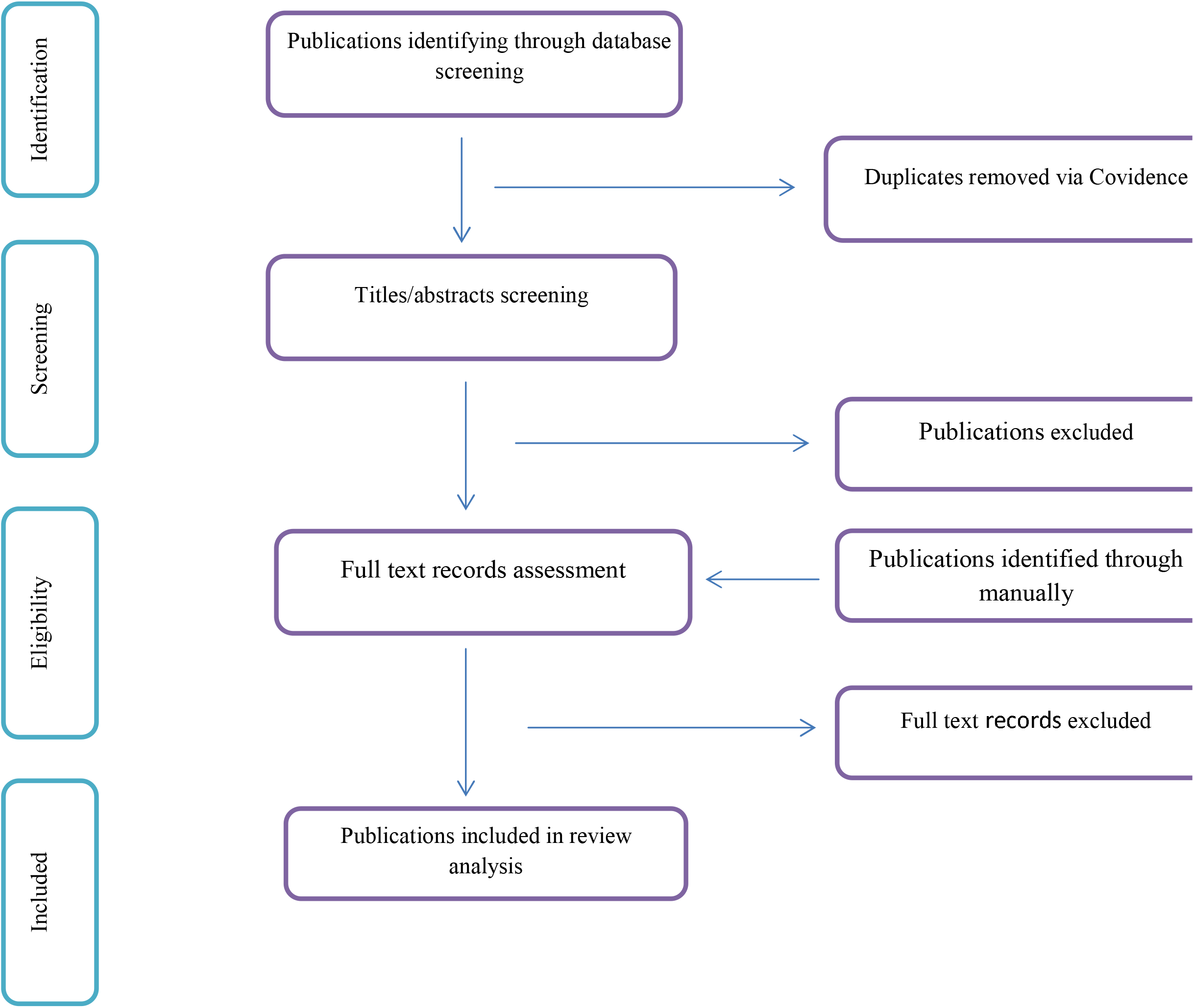
The PRISMA flow diagram

### 2.4. Charting the Data

Before commencing the data extraction phase, an extraction table will be crafted to align with the review’s objectives and research questions. Subsequently, a trial run of the extraction form will be conducted using a randomly selected sample of full-text records, encompassing five to eight records based on the number of studies included. Two reviewers will independently extract key information from the selected studies, meticulously recording their findings in a Microsoft Excel sheet. In the event of any discrepancies in the data extracted by the two team members, vibrant discussions will ensue. These spirited exchanges will continue until a harmonious consensus is reached, or the melodious input of the third reviewer will be sought for a harmonious resolution.

**Table 1.**
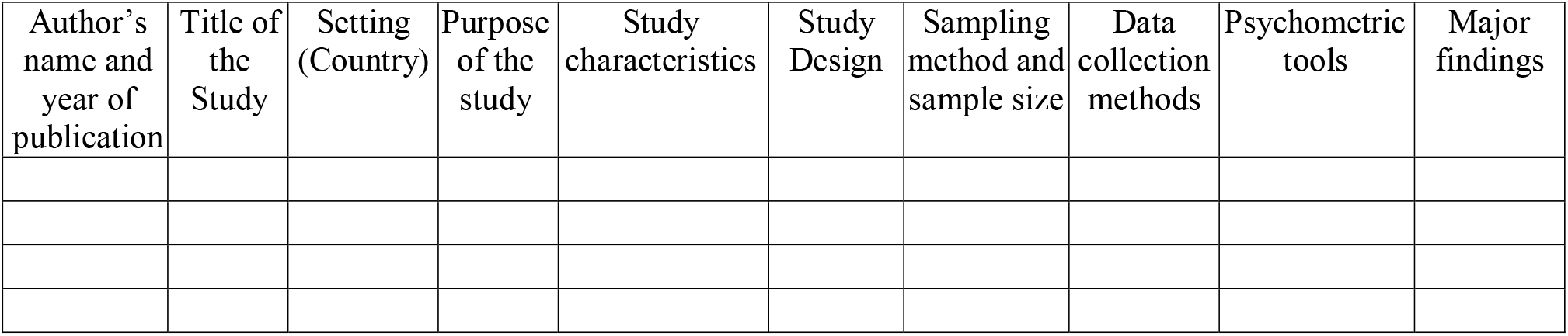
preliminary charting table.

### 2.5. Collating, summarizing, and reporting the findings

The selected records will be imported into NVivo Version 14, qualitative data analysis software. Two reviewers will independently code a 10% random sample of the records. This initial assessment aims to compute Cohen’s kappa coefficient, a statistical measure for assessing inter-rater reliability without necessitating discussion. A Cohen’s kappa coefficient exceeding 0.80 indicates a picture of satisfactory reliability, setting the stage for insightful findings (Thomas et al., 2017).

Once consensus is reached, the remaining selected records will be independently coded by each reviewer. To ensure consistency and reliability, regular meetings will be scheduled between the two coders. These meetings will facilitate the comparison of coding results, the resolution of any coding discrepancies, and the review of the codebook’s content.

Where applicable, quantitative data will be visually represented through diagrams, tables, or other descriptive formats. Concurrently, qualitative findings will be subjected to thematic analysis to identify key themes. In the final phase, the major findings will be reported in alignment with the objectives of this scoping review. Recognizing the inherent flexibility of such reviews, adjustments to the approach and procedures will be made as deemed necessary

## 3. Ethics and Dissemination

Instead of assessing the quality of each study individually, the report will deliver a thorough overview of existing evidence, consolidate findings, and present a summary outline. Given that the review relies on publicly accessible publications and materials, ethical approval is not necessary.

Following the completion of our review, this information will be disseminated among health administrators, relevant professionals, and researchers through publication in high-impact journals and presentation at conferences. Moreover, the gaps identified in the review will be instrumental in informing the principal investigator’s dissertation proposal.

## 4. Strength and Limitations

As far as our understanding extends, this scoping review will be the inaugural attempt to systematically explore, map, and pinpoint research gaps, thus paving the way for future research endeavors concerning the social determinants of adolescent depression in LMICs. Moreover, it will furnish policymakers, researchers, and healthcare professionals with invaluable insights to craft targeted interventions.

Our method is not free from some limitations. It is crucial to understand that scoping reviews are primarily intended to map the scope and breadth of the literature, not to delve into its specifics, such assessing the findings’ validity. Moreover, rather than producing new knowledge, scoping studies aim to describe knowledge gaps in the literature. We will not examine research quality or bias, nor will we systematically assess the evidence’s external validity, as is common in systematic reviews. Rather, we will concentrate on summarizing the salient features of the best-available data regarding the ways in which the contexts of the home, school, and community either promote or inhibit the development of depressive symptoms.

The review will specifically target studies published from 2010 to the end of 2023, potentially overlooking older yet pertinent research. Moreover, our inclusion criteria were restricted to English language records, potentially excluding significant non-English publications. The availability and quality of data in the included studies may also place restrictions on the breadth of the review, which could affect the precision and thoroughness of our conclusions. Furthermore, while the study focuses on pertinent depressive symptoms in adolescents, it does not assess how therapies affect depression. As a result, the quality of every contained record was not evaluated using a conventional appraisal instrument.

However, in a study by Nussbaumer-Streit et al. (2018), it was suggested that false conclusions stemming from literature searches can be minimized by utilizing multiple electronic literature databases and reviewing at least 10 records during the process. With this review utilizing six electronic literature databases, we anticipate that our findings will be presented with confidence.

## Data Availability

This is a scoping study protocol

## Funding

This research was funded by the NIHR (NIHR133712) using UK international development funding from the UK Government to support global health research. The views expressed in this publication are those of the author(s) and not necessarily those of the NIHR or the UK government, the Court of the University of Aberdeen, the Board of Directors of the University of Rwanda, the Board of Directors of Addis Ababa University, the Board of Directors of The Sanctuary, or our International Advisory Board.

## Ethical and Institutional Review

Not applicable

## Consent for Publication

Not applicable

## Conflict of Interest

The authors declare that they have no competing interests.

## Notes

### Competing Interest Statement

The authors have declared no competing interest.

## References

Abela, J. R. Z., & Hankin, B. L. (2008). Depression in Children and Adolescents: Causes, Treatment, and Prevention The Guilford Press.

Alshammari, A. S., Piko, B. F., & Fitzpatrick, K. M. (2021). Social support and adolescent mental health and well-being among Jordanian students. International Journal of Adolescence and Youth, 26(1), 211–223. 10.1080/02673843.2021.1908375

Arksey, H., & O’Malley, L. (2005). Scoping studies: towards a methodological framework. International Journal of Social Research Methodology., 19–32. 10.1080/1364557032000119616

De-la-Iglesia, M., & Olivar, J. S. (2015). Risk Factors for Depression in Children and Adolescents with High Functioning Autism Spectrum Disorders. ScientificWorldJournal, 2015, 127853. 10.1155/2015/127853

Frojd, S. A., Nissinen, E. S., Pelkonen, M. U., Marttunen, M. J., Koivisto, A. M., & Kaltiala-Heino, R. (2008). Depression and school performance in middle adolescent boys and girls. J Adolesc, 31(4), 485–498. 10.1016/j.adolescence.2007.08.006

Hankin, B. L. (2006). Adolescent depression: Description, causes, and interventions. Epilepsy and Behavior, 8(1), 102–114. 10.1016/j.yebeh.2005.10.012

Hunduma, G., Deyessa, N., Dessie, Y., Geda, B., & Yadeta, T. A. (2022). High Social Capital is Associated with Decreased Mental Health Problem Among In-School Adolescents in Eastern Ethiopia: A Cross-Sectional Study. Psychol Res Behav Manag, 15, 503–516. 10.2147/PRBM.S347261

Ibrahim, N., Sherina Mohd Sidik, Phang Cheng Kar, Firdaus Mukhtar, Ramidin Awang, Ang Jin Kiat, Osman, Z. J., & Siti Fatimah Ab Ghaffar. (2017). Prevalence and predictors of depression and suicidal ideation among adolescents attending government secondary schools in Malaysia. The Medical journal of Malaysia, 72, No 4.

Jason, M. N., Sejal, H. B, Jane, F., Michele, J. H., Sachiyo, Y., & David, A. R. (2018). Research priorities for adolescent health in low- and middle-income countries: A mixed-methods synthesis of two separate exercises. J Glob Health, 8(1). 10.7189%2Fjogh.08.010501

Jayanthi, P., Thirunavukarasu, M., & Rajamanickam, R. (2015). Academic Stress and Depression among Adolescents: A Cross-sectional Study Indian Pediatrics, 52, 217–219.

Jenness, J. L., Peverill, M., King, K. M., Hankin, B. L., & McLaughlin, K. A. (2019). Dynamic associations between stressful life events and adolescent internalizing psychopathology in a multiwave longitudinal study. J Abnorm Psychol, 128(6), 596–609. 10.1037/abn0000450

McHugh, M. (2012). Interrater reliability: the kappa statistic. Biochem Med (Zagreb), 22(3), 276–282. https://www.ncbi.nlm.nih.gov/pmc/articles/PMC3900052/

Mekdes, D. B., Angaw, D. A., & Mulat, H. (2018). Prevalence and Associated Factors of Depression among Orphan Adolescents in Addis Ababa, Ethiopia. Psychiatry J, 2018, 5025143. 10.1155/2018/5025143

Minichil, W., Getinet, W., Derajew, H., & Seid, S. (2019). Depression and associated factors among primary caregivers of children and adolescents with mental illness in Addis Ababa, Ethiopia. BMC Psychiatry, 19(1), 249. 10.1186/s12888-019-2228-y

Mukangabire, P., Moreland, P., Kanazayire, C., Rutayisire, R., Nkurunziza, A., Musengimana, D., & Kagabo, I. (2021). Prevalence and Factors Related to Depression among Adolescents Living with HIV/AIDS, in Gasabo District, Rwanda. Rwanda Journal of Medicine and Health Sciences 4.

Muntaner, C., Edwin, N., Vanroelen, C., Sharon, C., & William, W. E. (2013). Social Stratification, Social Closure, and Social Class as Determinants of Mental Health Disparities In Handbook of the Sociology of Mental Health (pp. 299–323). 10.1007/978-94-007-4276-5_15

Nussbaumer-Streit, B., Klerings, I., Wagner, G., Heise, T. L., Dobrescu, A. I., Armijo-Olivo, S., Stratil, J. M., Persad, E., Lhachimi, S. K., Van Noord, M. G., Mittermayr, T., Zeeb, H., Hemkens, L., & Gartlehner, G. (2018). Abbreviated literature searches were viable alternatives to comprehensive searches: A meta-epidemiological study. Journal of Clinical Epidemiology, 102, 1–11. 10.1016/j.jclinepi.2018.05.022

Petersen, B.E. Compas, J. Brooks-Gunn, M. Stemmler, S.E., and, & K. E. Grant. (1993). Depression in Adolescence. American Psychologist, 48, no. 2, 155–168.

Presentati, J.-e. a., (2021). The prevalence of mental health problems in subsaharan adolescents: a systematic review. PLoS ONE., 16(5).

Rehm, J., & Shield, K. (2019). Global Burden of Disease and the Impact of Mental and Addictive Disorders. Curr Psychiatry Rep., 21(2). 10.1007/s11920-019-0997-0.

Schaan, V. v. B. (2013). Social Determinants of Depression in Later Life University of Mannheim].

Shefaly, S., Esperanza Debby Ng, & Wong, C. H. J. (2022). Global prevalence of depression and elevated depressive symptoms among adolescents: A systematic review and meta-analysis. British Journal of Clinical Psychology, 61, 287–305. 10.1111/bjc.12333

Sreekanth, P., Poojitha, M., Kumar, G. B., Ugandar, D. R. E., Setlem, V. S. K., Pottipadu, D. V., Molugulu, D. N., & Chinnappan, S. (2023). Prevalence and Coping Strategies of Depression, Anxiety and Stress among High School Adolescents: A Cross Sectional Study. Journal of Population and Therapeutics and Clinical Pharmacology. 10.53555/jptcp.v30i3.2508

Tirfeneh, E., & Srahbzu, M. (2020). Depression and Its Association with Parental Neglect among Adolescents at Governmental High Schools of Aksum Town, Tigray, Ethiopia, 2019: A Cross Sectional Study. Depression Research and Treatment, 2020, 1–9. 10.1155/2020/6841390

Turner, R. J., Turner, J. B., & William, B. H. (2014). Social Relationships and Social Support. In R. J. Johnson, T. Jay, & G. L. Bruce (Eds.), Sociology of Mental Health Selected Topics from Forty Years 1970s–2010s (pp 1–21). Springer.

Upashree D., & Maraichelvi, K. A. (2020). Stress, Anxiety and Depression among Adolescents Ethno Med, 14(1-2), 68–74 10.31901/24566772.2020/14.1-2.606

Vos, T., Allen, C., & Arora, M. (2016). Global, regional, and national incidence, prevalence, and years lived with disability for 310 diseases and injuries, 1990-2015: a systematic analysis for the Global Burden of Disease Study 2015. The Lancet, vol. 388, no. 10053,, pp. 1545–1602,

Wang, L., Feng, Z., Yang, G., Yang, Y., Wang, K., Dai, Q., Zhao, M., Hu, C., Zhang, R., Liu, K., Guang, Y., & Xia, F. (2016). Depressive symptoms among children and adolescents in western china: An epidemiological survey of prevalence and correlates. Psychiatry Res, 246, 267–274. 10.1016/j.psychres.2016.09.050

Weersing, V. R., Shamseddeen, W., Garber, J., Hollon, S. D., Clarke, G. N., Beardslee, W. R., Gladstone, T. R., Lynch, F. L., Porta, G., Iyengar, S., & Brent, D. A. (2016). Prevention of Depression in At-Risk Adolescents: Predictors and Moderators of Acute Effects. J Am Acad Child Adolesc Psychiatry, 55(3), 219–226. 10.1016/j.jaac.2015.12.015

WHO. (2017). Depression: let’s talk. https://www.who.int/news-room/events/detail/2017/04/07/default-calendar/world-health-day-2017

WHO. (2018b). Adolescents: health risks and solutions. World Health Organization.

WHO. (2021). Mental health of adolescents. Retrieved 12/12 from https://www.who.int/news-room/fact-sheets/detail/adolescent-mental-health

WHO. (2022). COVID-19 pandemic triggers 25% increase in prevalence of anxiety and depression worldwide. Retrieved 15/2 from https://www.who.int/news/item/02-03-2022-covid-19-pandemic-triggers-25-increase-in-prevalence-of-anxiety-and-depression-worldwide

Zhu, S., & Wong, P. W. C. (2022). What matters for adolescent suicidality: Depressive symptoms or fixed mindsets? Examination of cross-sectional and longitudinal associations between fixed mindsets and suicidal ideation. Suicide Life Threat Behav, 52(5), 932–942. 10.1111/sltb.12891

